# Use of low-dose decitabine with or without tyrosine kinase inhibitors in advanced phase chronic myelogenous leukemia: a systemic review and metaanalysis

**DOI:** 10.1101/2022.05.06.22274754

**Authors:** Maureen Via M. Comia, Charles Eryll S. Sy, Jomell C. Julian

**Author notes:** Corresponding author: Maureen Via M. Comia, MD, FPCP.

## Abstract

**Rationale:** Progression of Chronic Myelogenous Leukemia (CML) to more advanced phases can involve hypermethylation, which is correlated to resistance or intolerance to imatinib. This hypermethylation has also been found to be a negative prognostic factor independent of imatinib response and from CML phase, thus decitabine, a hypomethylating agent, can be an attractive treatment option for advanced phase CML.

**Objective:** This systemic review and meta-analysis aims to investigate the role of low-dose decitabine among patients with advanced phase CML.

**Methodology:** This was performed according to the statement of Preferred Reporting Items for Systematic Reviews and Meta-Analyses (PRISMA)

**Results:** Four (4) studies from 86 articles screened were eligible to be assessed in this systemic review and meta-analysis. These were phase I/II trials involving 81 advanced phase CML patients and used low-dose decitabine (5 to 20 mg/m^2^), with two studies using tyrosine kinase inhibitors. Outcomes of hematologic and cytogenetic response, and survival were assessed in the meta-analysis; with hematologic response being favored among advanced phase CML patients upon exposure with low-dose decitabine *(p=0*.*05)*. Survival was also favored among responders to low-dose decitabine, however this was not significant.

**Discussion and Conclusion:** Low-dose decitabine can be an effective and safe treatment option in advanced phase CML, especially in more frail patients that could not tolerate more intensive chemotherapy regimens. However, this study is limited by few studies available on this topic, thus further randomized controlled trials can be investigated to define the role of decitabine and its optimal dose among this subset of patients.

## INTRODUCTION

Chronic Myelogenous Leukemia (CML) is a myeloproliferative neoplasm, characterized by the reciprocal chromosomal translocation between the long arms of chromosome 9 and 22 [(t9;22)(q34;11)], resulting to a shortened chromosome 22, also known as the Philadelphia chromosome.^1,2^ The resultant fusion oncogene BCR/ABL1 encodes a constitutive active yet defective tyrosine kinase, which is a pathogenic driver capable of initiating and maintaining the disease.^1,3^

CML has a worldwide annual incidence rate of 0.87 to 1.52 per 100,000, and this incidence increases with age. Median age of diagnosis is 56 years old, with slight male predominance.^1^ Clinical manifestations range from asymptomatic, as diagnosed incidentally on routine complete blood count, to symptoms related to anemia and splenomegaly. These symptoms are mostly seen among patients with chronic phase CML, with almost 90% of patients being diagnosed in this phase.^1,3^ Symptoms from hyperleukocytosis and hyperviscosity, such as priapism, tinnitus, or stupor, can also be seen. Chronic phase CML can progress to accelerated and blastic phases, which manifest more worrisome symptoms such as fever, bone and joint pains, bleeding, infections and lymphadenopathy. Accelerated phase CML can be defined by a set of criteria developed by MD Anderson Cancer Center (MDACC) and involves presence of abnormal blood counts and additional clonal cytogenetic abnormalities. A definition of blastic phase CML was also given by International Bone Marrow Registry and involves presence of ≥30% blasts in peripheral blood or bone marrow, or both, or presence of extramedullary infiltrates of leukemic cells.^4^

Development of advanced phase CML from chronic phase has been extensively studied. In a study by Bavaro et al. (2019), they described that progression from chronic phase to more advanced phase involves block of differentiation and apoptosis, alterations in cell adhesion, activation of alternative signaling pathways, and a shift toward turning on expression of genes involved in the nucleosome. BCR/ABL1 was also observed to increase as the disease progresses, which then promotes onset of secondary molecular and chromosomal hits, leading to expansion of highly proliferating differentiation-arrested malignant cell clones. However, when these hits have been acquired, inhibiting BCR/ABL1 alone often fails, as manifested by progression even with tyrosine kinase inhibitor (TKI) treatment. This shows that there are other several BCR/ABL1 independent mechanisms involved in progression of CML to advanced phase.^3^ These findings were also reflected on previous studies of Shet et al. (2002) and Callabretta and Perrotti (2004).^5,6^ Bavaro et al. (2019) also described the methylation changes causing progression of chronic phase CML to advanced phase CML. This hypermethylation has been correlated to imatinib resistance or intolerance and found to be a negative prognostic factor independent of imatinib response and from CML phase.^3^

This finding led to clinical trials on use of a hypomethylating agent, decitabine, on advanced phase CML. Decitabine also has several other properties such as differentiation induction, anti-leukemic efficacy and synergism with interferons and retinoids^7^ thus, making it an attractive treatment option for a seemingly hard-to-treat advanced phase CML. This systematic review and meta-analysis aims to concise and investigate the evidences regarding the efficacy of decitabine in advanced phase CML, and assess its impact on our treatment decision-making in these kinds of patients.

## OBJECTIVES

### General Objective

This systemic review and meta-analysis aims to investigate the role of low-dose decitabine among patients with advanced phase chronic myelogenous leukemia.

### Specific Objectives

1. To determine efficacy of low-dose decitabine in terms of hematologic response among patients with advanced phase CML;
2. To determine efficacy of low-dose decitabine in terms of cytogenetic response among patients with advanced phase CML; and,
3. To determine efficacy of low-dose decitabine in terms of survival among patients with advanced phase CML.

### DEFINITION OF TERMS

1. Advanced phase chronic myelogenous leukemia – CML that has progressed to accelerated or blastic phase
2. Accelerated phase chronic myelogenous leukemia – CML with peripheral blast of 10-19%, peripheral blood basophils >20%, thrombocytopenia of <100 × 10^9^/L unrelated to therapy and new clonal cytogenetic abnormalities accompanying the Philadelphia chromosome
3. Blastic phase – CML evolve to overt acute leukemia, either myeloid or lymphoid
4. Complete hematologic response – complete normalization of peripheral blood counts with leukocyte count of <10 × 10^9^/L
5. Complete cytogenetic response – no Philadelphia chromosome-positive metaphases
6. Molecular response – MR4.5 (BCR/ABL1 ratio <0.0032% international scale [IS])
7. Low-dose decitabine – described in several clinical trials as 5-20 mg/m^2^/day

## METHODS

### Study Design

This systemic review and meta-analysis was performed according to the statement of Preferred Reporting Items for Systematic Reviews and Meta-Analyses (PRISMA). The two authors independently performed the literature search, evaluated study eligibility, extracted the relevant data, and assessed the risk of bias of each study. Discrepancies were resolved through discussion and consultation with the third author.

### Eligibility Criteria (Inclusion and Exclusion)

Published randomized controlled trials and non-randomized studies, either prospective or retrospective, were eligible for inclusion with no minimum number of patients. The study population consisted of patients diagnosed with advanced phase chronic myelogenous leukemia receiving low-dose decitabine chemotherapy with or without tyrosine kinase inhibitors (TKIs). The primary outcomes measured were hematologic and cytogenetic response, and survival. Secondary outcomes were molecular response and adverse events.

There was exclusion of publications written in a language other than English, review papers, and on-going clinical trials. There were no restrictions on sex, ethnicity or clinical setting.

### Search Strategies

A systematic search of the databases was conducted, utilizing the PubMed and Cochrane Library to identify relevant published literature to address the research objective with a deadline of April 2022. The search terms used were “chronic myelogenous leukemia” OR “CML” AND “decitabine”. Bibliographies of relevant studies identified were searched for additional material and authors.

### Data Collection and Analysis

#### Selection of Studies

The study selection process was conducted following the PRISMA criteria. After the removal of duplications, articles were screened based on the inclusion and exclusion criteria. The two authors independently screened the results of the search strategies for the eligibility by reading the abstracts.

Following this, the two authors assessed independently the full-text articles of selected studies. Discrepancies were resolved through discussion and consultation with the third author.

### Data Extraction and Management

The eligible studies were reviewed in full-text independently by the authors. Methodological quality and risk of bias assessment were done for each included study. The data extracted from the included studies were article title, name of the author(s), date of publication, study design, methodological features (randomization, allocation concealment, blinding measures), study population, participant characteristics, TKI used, number of cycles and dose of decitabine, and outcomes (hematologic, cytogenetic, and molecular response, survival, and adverse events). The data obtained were summarized using Microsoft Excel. All data were compared for consistency.

### Risk of Bias Assessment

The risk of bias of non-controlled non-randomized studies of interventions were assessed using the risk of bias assessment criteria for observational studies tool provided by Cochrane Childhood Cancer. This tool includes six important domains that should be considered: selection bias, attrition bias, detection bias, confounding bias, reporting bias and analysis bias. Each of the domains was judged as low, unclear or high risk of bias, and an overall grade for the risk of bias will be concluded as well.

### Data Synthesis and Assessment of Heterogeneity

For data analysis, descriptive statistics were used to summarize the baseline characteristics. The type of outcome is dichotomous for hematologic and cytogenetic response, and survival. The statistical method used was Mantel-Haenszel method, with effect measure of odds ratio for hematologic and cytogenetic response, while Inverse Variance method was used for survival, with effect measure of hazard ratio. Forest plots, the chi-square test for heterogeneity, and the I2 statistic assessed statistical heterogeneity between studies. Funnel plot/Begg’s test was used to evaluate publication bias. Statistical significance was set at 0.05. All statistical analyses were performed using Revman version 5.4.1. Where meta-analysis was not feasible, a narrative synthesis was provided instead.

### Assessment of the Certainty of the Evidence

The authors used the Grading of Recommendations, Assessment, Development and Evaluations (GRADE) tool to assess the certainty of the evidence. GRADE identified its five categories: risk of bias, imprecision, inconsistency, indirectness, and publication bias. The certainty of evidence for non-randomized studies will start from low-certainty evidence.

## RESULTS

### Study Selection

The literature search yielded 82 articles from PubMed and four (4) from the Cochrane library. Four studies were excluded for duplicates. Twelve studies were screened based on their titles and abstracts. After reviewing the articles, four publications did not meet the study objective and were excluded. A total of four studies from the selection process were included in this study as illustrated in Figure 1. Full-text copies of the included studies were obtained for a more detailed examination.

**Figure 1.**
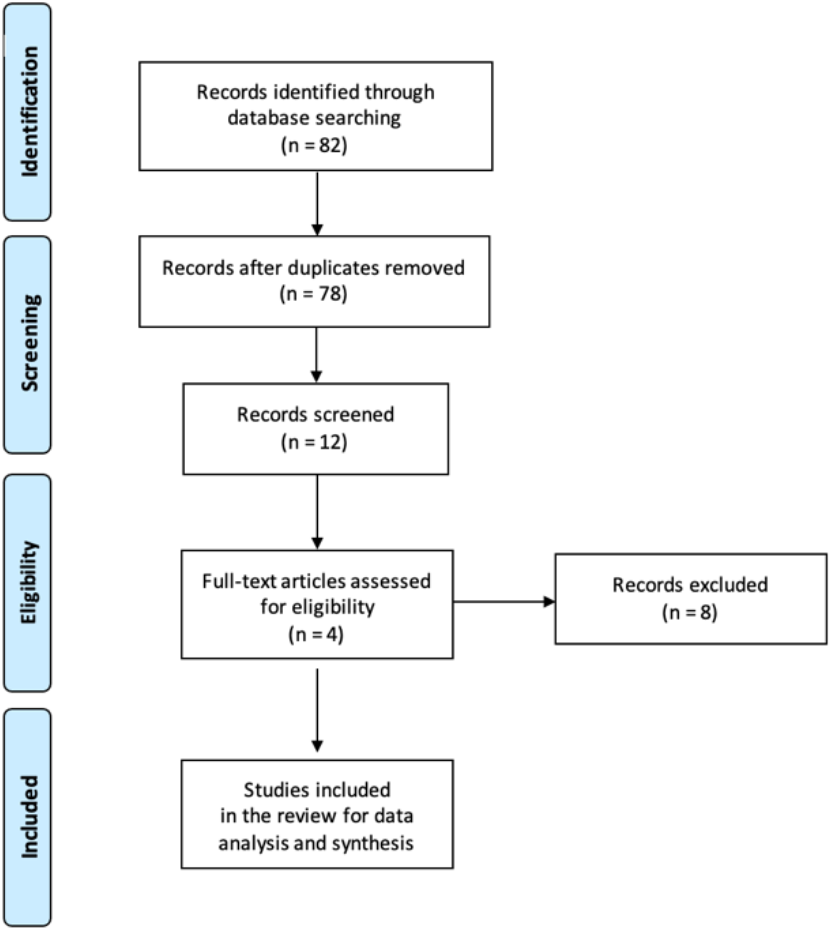
Study Selection

### Study Characteristics

The four articles included were published between 2004 and 2020. A total of 81 advanced phase chronic myelogenous leukemia adult patients were included in this review. The characteristics of the studies included in this review are summarized in Table 1. All included studies were published in English, and all were either phase I or phase II trials. Decitabine doses vary among the included studies ranging from 10 mg/m^2^ to 15 mg/m^2^ IV for 10 days.^8-11^

**Table 1.** General Characteristics of Included Studies

| **Table 1. General Characteristics of Included Studies** |  |  |  |  |  |  |
| --- | --- | --- | --- | --- | --- | --- |
| **Record No.** | **Author(s)** | **Date Published** | **Study Design** | **CML (n)** | **Decitabine dose** | **TKI used** |
| 1 | Issa et al. | March 2004 | Phase I trial | 4 | 15 mg/m<sup>2</sup> IV for 10 days | NR |
| 2 | Issa et al. | March 2005 | Phase II trial | 23 | 15 mg/m<sup>2</sup> IV for 10 days | NR |
| 3 | Oki et al. | November 2006 | Phase II trial | 28 | 15 mg/m<sup>2</sup> IV for 10 days | Imatinib 600 mg daily |
| 4 | Abaza et al. | July 2020 | Phase I/II trial | 26 | 10 mg/m<sup>2</sup> IV for 10 days | Dasatinib 100 mg or 140 mg daily |
| *CML = Chronic Myelogenous Leukemia; TKI = Tyrosine Kinase Inhibitors; NR = Not Reported* |  |  |  |  |  |  |

Patient characteristics, their reported responses with low-dose decitabine, median survival in weeks for both responders and non-responders, and overall number of patients with reported adverse events, for both hematologic and non-hematologic, are all summarized in tables 2, 3 and 4, respectively, for all the included studies.

**Table 2.** Patient Characteristics and Response in Included Studies

| **Table 2. Patient Characteristics and Response in Included Studies** |  |  |  |  |  |  |  |
| --- | --- | --- | --- | --- | --- | --- | --- |
| **Study** | **CML (n)** |  | **Median age in years** | **Sex Predominance** | **Overall Hematologic Response (%)** | **Overall Cytogenetic Response (%)** | **Molecular Response (%)** |
|  | **AP** | **BP** |  |  |  |  |  |
| Issa 2004 | 1 | 3 | 60 | Male | 40 | NR | NR |
| Issa 2005 | 17 | 6 | 61 | Male | 59/50 | 41/33 | NR |
| Oki 2006 | 18 | 10 | 50 | Female | 50/30 | 6/2 | NR |
| Abaza 2020 | 7 | 19 | 51 | Male | 83 | 52 | 33 |
| *CML = Chronic Myelogenous Leukemia; AP = Accelerated Phase; BP = Blastic Phase; NR = Not Reported* |  |  |  |  |  |  |  |

**Table 3.** Median Survival in Weeks

| **Study** | **Median survival in weeks, Responders** | **Median survival in weeks, Non-responders** |
| --- | --- | --- |
| Issa 2004 | 24 | NR |
| Issa 2005 | 8 | 18 |
| Oki 2006 | 86 | 16 |
| Abaza 2020 | 55.2 | 18.6 |
| *NR = Not Reported* |  |  |

**Table 4.** Overall Number of Patients with Reported Adverse Events in Included Studies

| **Study** | **Hematologic AE** | **Non-hematologic AE** |
| --- | --- | --- |
| Issa 2004 | N/R | 28 |
| Issa 2005 | 18 | N/R |
| Oki 2006 | 19 | 20 |
| Abaza 2020 | N/R | 26 |
| *AE = Adverse Event; NR = Not Reported* |  |  |

### Risk of Bias in Included Studies

Using the risk of bias assessment criteria for observational studies tool provided by Cochrane Childhood Cancer, the authors judged the overall risk of bias within and across the studies to be moderate. The full judgment for the studies is presented in Table 5.

**Table 5.** Risk of Bias Assessment

| **Domain** | **Issa 2004** | **Issa 2005** | **Oki 2006** | **Abaza 2020** |
| --- | --- | --- | --- | --- |
| **Selection bias** | Low | Low | Low | Low |
| **Attrition bias** | Low | Low | Low | Low |
| **Detection bias** | Low | Low | Low | Low |
| **Confounding bias** | Low | Low | Low | Low |
| **Reporting bias** | Low | Low | Low | Low |
| **Analysis bias** | Unclear | Unclear | Unclear | Unclear |
| **Overall risk of bias** | Moderate | Moderate | Moderate | Moderate |

## META-ANALYSIS

Primary outcomes measured in this metanalysis are hematologic and cytogenetic response, and survival.

Figure 2 shows the forest plot describing the hematologic response as assessed among all the patients included in the four studies. All four studies are non-randomized, prospective studies; and included patients were compared based on presence of overall hematologic response, including complete and partial hematologic response, and hematologic improvement, and non-response. Odds ratio was computed between the two groups on all the studies, with confidence interval of 95%. As seen in the forest plot, three out of four studies have an increased frequency of overall hematologic response upon exposure with low-dose decitabine. Overall estimated effect was also statistically significant, favoring hematologic response among advanced phase CML patients upon exposure with low-dose decitabine.

**Figure 2.**
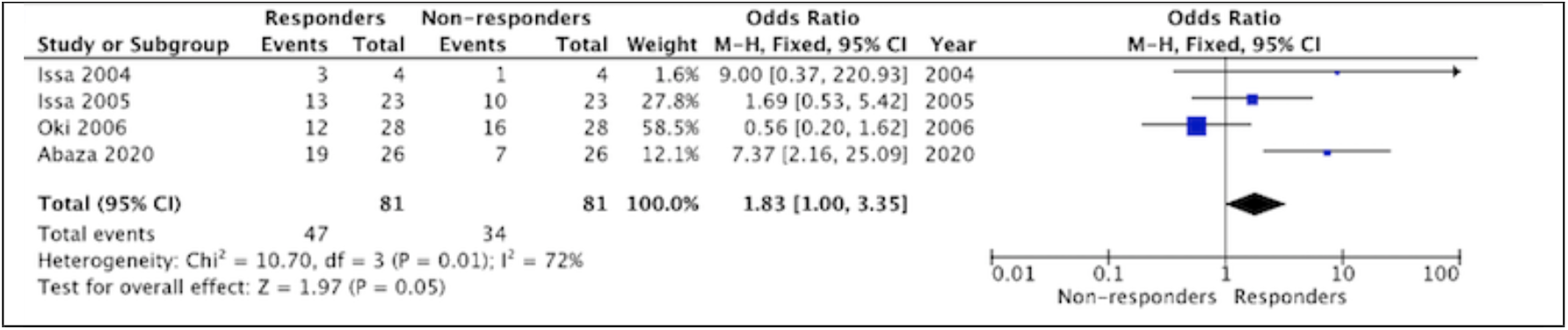
Hematologic Response of Advanced Phase CML Patients with Low-Dose Decitabine with or without TKI

Heterogeneity is also described in this forest plot. On Chi-squared test, p value was less than 0.1%, thus homogeneity among studies can be assumed; however, I2 statistic estimated all four studies with substantial heterogeneity. This could be explained by the nature of the studies being non-randomized, and can probably be overcomed by doing a subgroup analysis among the hematologic responses of these patients.

Figure 3 is the funnel plot for the outcome of hematologic response, and has fixed effect summary estimate at 1.95. Standard error is approaching zero, with three studies being high-powered than one study. This could be caused by a smaller sample size on this study. However, this funnel plot is asymmetric, and probably due to the substantial heterogeneity seen in these studies, and not necessarily due to any publication bias. An objective test for funnel plot asymmetry can be done aside from an eyeball test.

**Figure 3.**
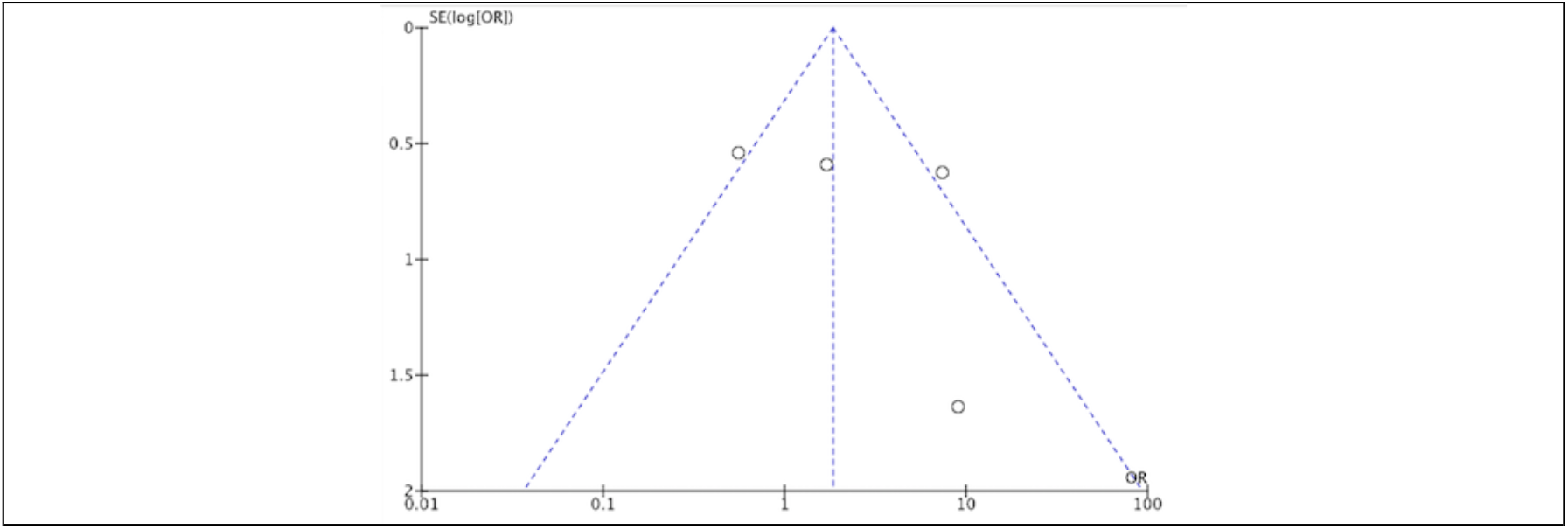
Hematologic Response of Advanced Phase CML Patients with Low-Dose Decitabine with or without TKI

Figure 4 shows the forest plot for cytogenetic response among the included patients. Only three out of four studies reported this outcome. As the graph is showing, cytogenetic response is not favored, and frequency of non-response is significantly more predominant at p value of 0.003. Heterogeneity is moderate amongst the studies; and could be explained by the nature of the studies being non-randomized, and could probably be overcomed with subgroup analysis.

**Figure 4.**
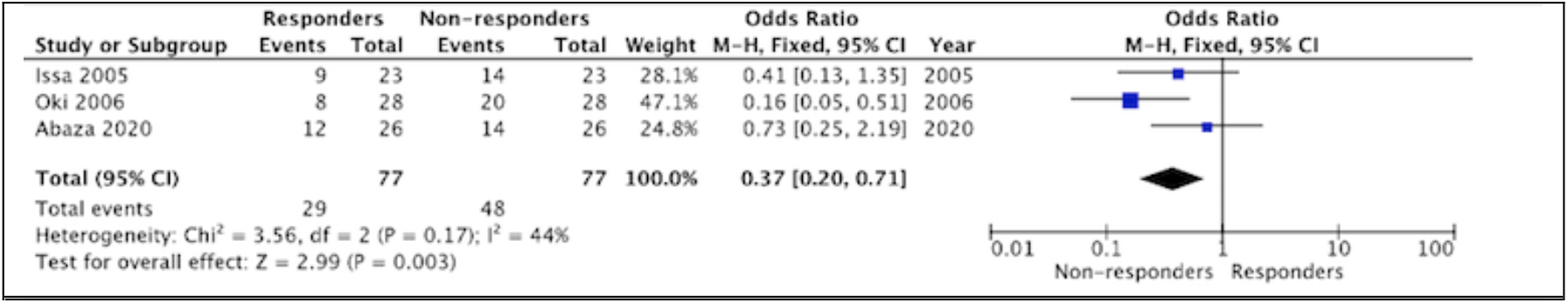
Cytogenetic Response of Advanced Phase CML Patients with Low-Dose Decitabine with or without TKI

Figure 5 is the funnel plot for cytogenetic response, and is symmetric with standard error approaching to zero. Included studies are almost of equal power, with no publication bias detected.

**Figure 5.**
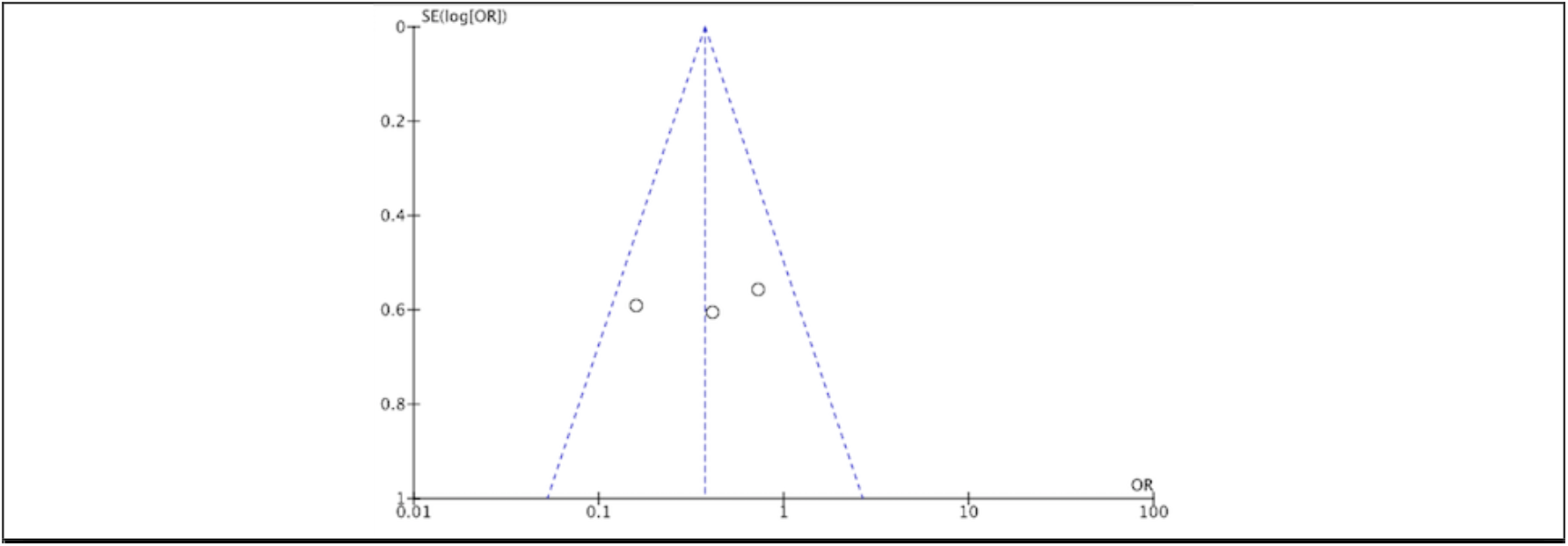
Cytogenetic Response of Advanced Phase CML Patients with Low-Dose Decitabine with or without TKI

Survival outcome was measured via hazard ratio. Not all four studies were included, as one study did not report survival outcome. Figure 6 shows the forest plot and describes that survival events can be seen more among responders to low-dose decitabine versus non-responders, with negative log hazard ratio indicating that there is decreased hazard and increased survival times among these responders. However, this is non-significant with a p value of 0.12. Heterogeneity upon I2 statistic is zero, concluding that the included studies are homogenous with each other.

**Figure 6.**
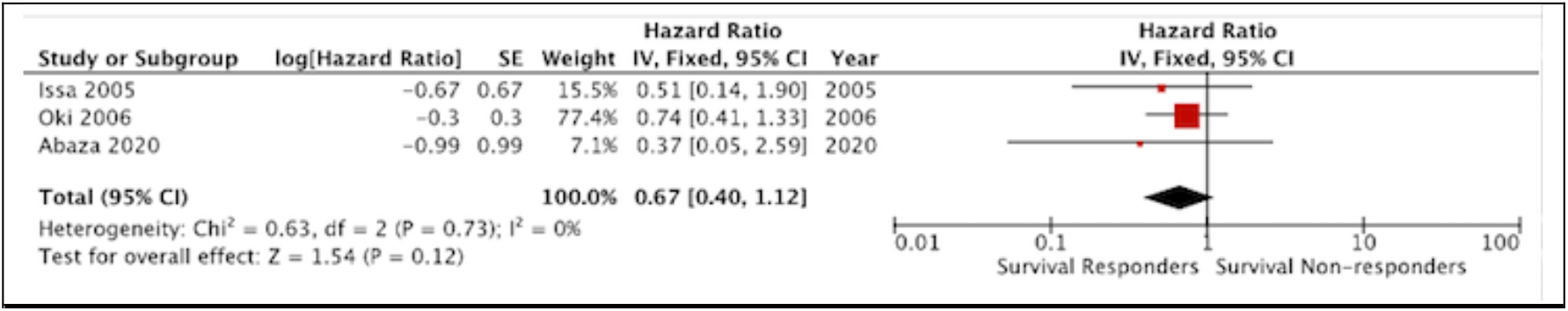
Survival of Advanced Phase CML Patients with Low-Dose Decitabine with or without TKI

Figure 7 is the funnel plot for survival, and is symmetric with standard error approaching to zero. Included studies are scattered according to power, with no publication bias detected.

**Figure 7.**
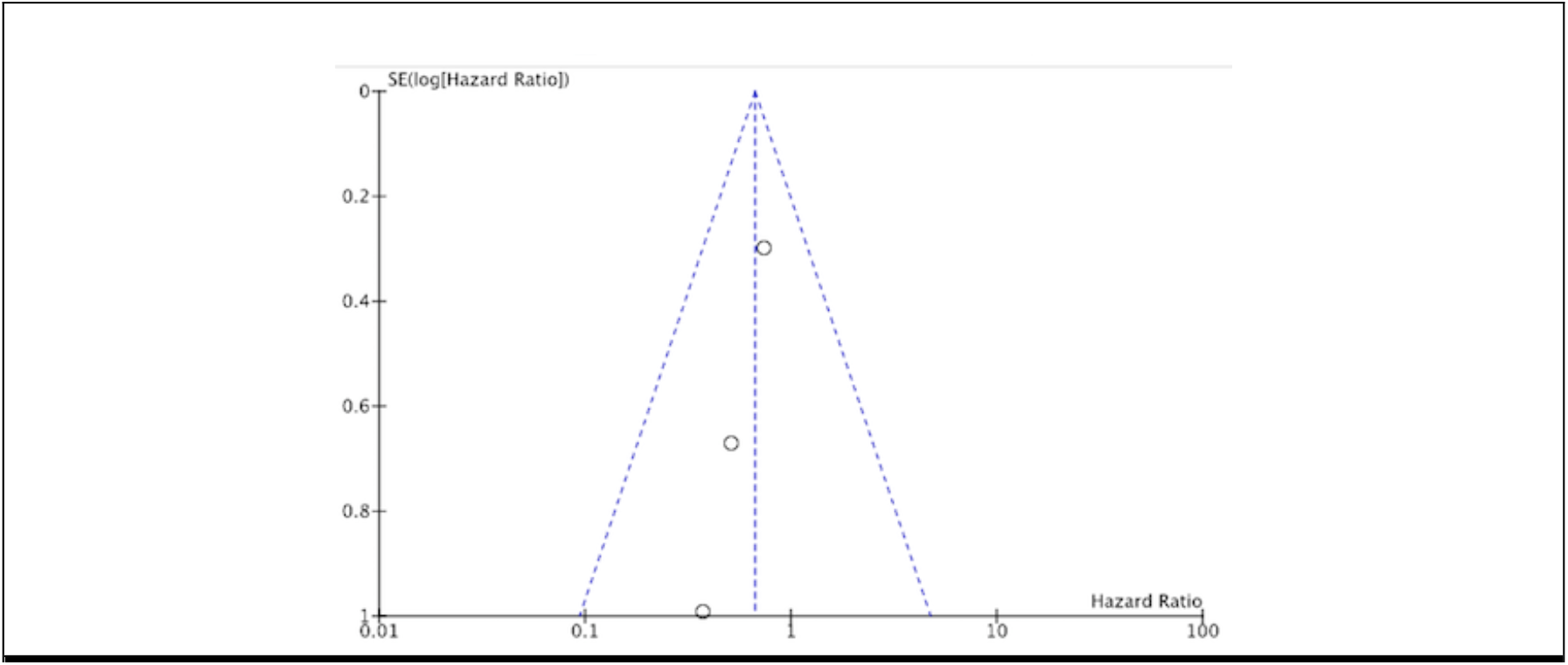
Survival of Advanced Phase CML Patients with Low-Dose Decitabine with or without TKI

### Certainty of Evidence

We used the GRADE approach to assess the certainty of the evidence for the following outcomes: hematologic response and survival. We assessed the certainty of evidence for non-controlled non-randomized studies of intervention starting from low-certainty evidence. The certainty of evidence in the reported outcomes was further reduced to very low because of the very small information size and moderate risk of bias of included studies.

## DISCUSSION

Chronic Myelogenous Leukemia (CML) in advanced phase has been traditionally challenging to manage as the disease can develop resistance towards TKIs as it progresses, and sometimes with addition of a more aggressive chromosomal aberrancy. The first generation TKI, Imatinib, can still be used in advanced phase CML, with response ranging from as low as 0% in blastic phase to as high as 90% in accelerated phase. Survival is seen at 50 – 60% at 5 years. Higher generations of TKIs, particularly Bosutinib and Ponatinib, are preferred and used for advanced phase CML, especially after Imatinib failure. Response ranges from 11% to 57%, with survival rate ranging from 60% at 4 years for Bosutinib to 84% at 1 year for Ponatinib.^12^ These responses were also reflected in an earlier study done by Bonifacio et al. (2019).^13^ However, other treatment modalities, like chemotherapy, and if possible, hematopoietic stem cell transplantation (HSCT), are generally recommended in advanced phase CML, especially in blastic phase. Several chemotherapy regimen options were studied in small, retrospective studies and included cytarabine-based regimens (7+3 and FLAG-Ida) for myeloid blast crisis, and Hyper-CVAD with Dasatinib in lymphoid blast crisis. These regimens produced a superior survival rates compared to TKI alone.^12^

Based on one of the mechanisms of disease progression in CML, decitabine has been explored even in the pre-TKI era, to address hypermethylation seen in advanced phase CML.^3^ In an earlier study by Kantarjian et al. (1997), 37 patients with advanced phase CML were treated with 75 to 100 mg/m^2^ decitabine for 10 doses. An overall response rate of 53% for accelerated phase was seen, with a lower rate at 25% for blastic phase. Prolonged myelosuppression was seen to be the most significant side effect.^7^ These modest results were reflected in follow-up studies done by Kantarjian et al. (2003)^14^ together with Sacchi et al. (1999)^15^, and listed better survival among patients treated with decitabine. These studies, however, used much higher doses as compared to the studies included in this meta-analysis. It produced prolonged and severe myelosuppression that can affect survival among these patients. In a study by Issa and Byrd (2005), they summarized studies using a much lower dose of decitabine, given in prolonged exposure schedule.^16^ These studies showed a significantly fewer responses in patients treated with higher doses, and lower doses were more tolerated. Issa et al. (2004)^8^ described in his study that low-dose decitabine affects silencing by aberrant methylation and normalizes the gene expression profile of malignant cells, while high dose decitabine makes DNA adducts eventually resulting in cytotoxicity, explaining the prolonged myelosuppression in high doses.^9^

With regards to overall hematologic response, studies included in this meta-analysis favored hematologic response and produced almost similar modest results (40% to 59%) with low-dose decitabine among accelerated phase CML patients as compared to higher doses. Higher responses (30% to 50%), however, were seen among blastic phase CML patients given low-dose decitabine compared to higher doses. An advantage of more tolerable myelosuppression was also seen. In the study by Abaza et al. (2020), a much higher overall hematologic response (56% to 83%) for accelerated and blastic phases was seen.^11^ This might be attributed to the use of Dasatinib among these patients, owing the better response to the synergy of hypomethylating agents and TKIs.

Cytogenetic responses were also investigated in most of the included studies. Total cytogenetic responses among these studies range from 2% to 52%. When compared to higher-dose decitabine studies, higher-dose studies reported lower cytogenetic response rates (0% to 9%). However, this outcome was seen to be not favorable towards response in this meta-analysis. Abaza et al. (2020)^11^ discussed that attaining cytogenetic and molecular response are low even with combination therapy, and could be seen more among patients that are in chronic phase.

Survival, meanwhile, was favored among responders to low-dose decitabine in this meta-analysis. Median survival in weeks among responders in these studies was as high as 86 weeks, compared to 18.6 weeks in non-responders. Higher-dose decitabine, meanwhile, also showed better survival amongst the patients, but rates were more modest as compared to those given with low-dose decitabine, owing to a more significant myelosuppression seen in higher-dose decitabine. Furthermore, these studies were mostly done in the pre-TKI era.

Aside from myelosuppression, other minor adverse events were reported in the included studies. Generally, treatment with low-dose decitabine was well tolerated. Non-severe adverse events include nausea, vomiting, diarrhea, mucositis, skin rashes, and mild elevations in liver enzymes and creatinine. Not considering the severe myelosuppression in higher-doses, decitabine can be considered as a safe treatment option, especially if given in low-dose.

## LIMITATIONS

There are several limitations in this meta-analysis to be considered. There is a limited number of related studies, and most had few sample sizes. Most of the studies included were performed before or at the early years of TKI treatment; hence some participants were not given TKI as part of their treatment, which may had affected their outcomes and survival.

## CONCLUSIONS

### Implications for Practice

There are limited options on effective treatment options for advanced phase CML, however this meta-analysis shows that low-dose decitabine can be an effective and safe treatment option, especially in more frail patients that could not tolerate more intensive chemotherapy regimens.

### Implications for Research

Only few studies were available regarding this topic. Further randomized controlled trials can be investigated to define the role of decitabine and its optimal dose among this subset of patients.

## Data Availability

All data produced in the present study are available upon reasonable request to the authors and contained in the manuscript.

## REFERENCES

1. Eden RE, Coviello JM. Chronic Myelogenous Leukemia. [Updated 2022 Jan 24]. In: StatPearls [Internet]. Treasure Island (FL): StatPearls Publishing; 2022 Jan-. Available from: https://www.ncbi.nlm.nih.gov/books/NBK531459/

2. Salesse, S., Verfaillie, C. BCR/ABL: from molecular mechanisms of leukemia induction to treatment of chronic myelogenous leukemia. Oncogene 21, 8547–8559 (2002). https://doi.org/10.1038/sj.onc.1206082

3. Bavaro L, Martelli M, Cavo M, Soverini S. Mechanisms of Disease Progression and Resistance to Tyrosine Kinase Inhibitor Therapy in Chronic Myeloid Leukemia: An Update. International Journal of Molecular Sciences. 2019; 20(24):6141. https://doi.org/10.3390/ijms20246141

4. National Comprehensive Cancer Network Clinical Practice Guidelines Chronic Myeloid Leukemia. Version 1.2022

5. Shet AS, Jahagirdar BN, Verfaillie CM. Chronic myelogenous leukemia: mechanisms underlying disease progression. Leukemia. 2002 Aug;16(8):1402–11. doi: 10.1038/sj.leu.2402577. PMID: 12145676.

6. Calabretta B, Perrotti D. The biology of CML blast crisis. Blood. 2004 Jun 1;103(11):4010–22. doi: 10.1182/blood-2003-12-4111. Epub 2004 Feb 24. PMID: 14982876.

7. Kantarjian, H., O’Brien, S., Keating, M. et al. Results of decitabine therapy in the accelerated and blastic phases of chronic myelogenous leukemia. Leukemia 11, 1617–1620 (1997). https://doi.org/10.1038/sj.leu.2400796

8. Issa, J. P. J., Garcia-Manero, G., Giles, F. J., Mannari, R., Thomas, D., Faderl, S., Bayar, E., Lyons, J., Rosenfeld, C. S., Cortes, J., & Kantarjian, H. M. (2004). Phase 1 study of low-dose prolonged exposure schedules of the hypomethylating agent 5-aza-2′-deoxycytidine (decitabine) in hematopoietic malignancies. Blood, 103(5), 1635–1640. https://doi.org/10.1182/blood-2003-03-0687

9. Issa JP, Gharibyan V, Cortes J, Jelinek J, Morris G, Verstovsek S, Talpaz M, Garcia-Manero G, Kantarjian HM. Phase II study of low-dose decitabine in patients with chronic myelogenous leukemia resistant to imatinib mesylate. J Clin Oncol. 2005 Jun 10;23(17):3948–56. doi: 10.1200/JCO.2005.11.981. Epub 2005 May 9. PMID: 15883410.

10. Oki, Y., Kantarjian, H. M., Gharibyan, V., Jones, D., O’Brien, S., Verstovsek, S., Cortes, J., Morris, G. M., Garcia-Manero, G., & Issa, J. P. J. (2007). Phase II study of low-dose decitabine in combination with imatinib mesylate in patients with accelerated or myeloid blastic phase of chronic myelogenous leukemia. Cancer, 109(5), 899–906. https://doi.org/10.1002/cncr.22470

11. Abaza, Y., Kantarjian, H., Alwash, Y., Borthakur, G., Champlin, R., Kadia, T., Garcia-Manero, G., Daver, N., Ravandi, F., Verstovsek, S., Burger, J., Estrov, Z., Ohanian, M., Lim, M., Pemmaraju, N., Jabbour, E., & Cortes, J. (2020). Phase I/II study of dasatinib in combination with decitabine in patients with accelerated or blast phase chronic myeloid leukemia. American Journal of Hematology, 95(11), 1288–1295. https://doi.org/10.1002/ajh.25939

12. How, J., Venkataraman, V., & Hobbs, G. S. (2021). Blast and accelerated phase CML: room for improvement. Hematology, 2021(1), 122–128. https://doi.org/10.1182/hematology.2021000240

13. Bonifacio, M., Stagno, F., Scaffidi, L., Krampera, M., & di Raimondo, F. (2019). Management of Chronic Myeloid Leukemia in Advanced Phase. Frontiers in Oncology, 9. https://doi.org/10.3389/fonc.2019.01132

14. Kantarjian HM, O’Brien S, Cortes J, Giles FJ, Faderl S, Issa JP, Garcia-Manero G, Rios MB, Shan J, Andreeff M, Keating M, Talpaz M. Results of decitabine (5-aza-2’deoxycytidine) therapy in 130 patients with chronic myelogenous leukemia. Cancer. 2003 Aug 1;98(3):522–8. doi: 10.1002/cncr.11543. PMID: 12879469.

15. Sacchi S, Kantarjian HM, O’Brien S, Cortes J, Rios MB, Giles FJ, Beran M, Koller CA, Keating MJ, Talpaz M. Chronic myelogenous leukemia in nonlymphoid blastic phase: analysis of the results of first salvage therapy with three different treatment approaches for 162 patients. Cancer. 1999 Dec 15;86(12):2632–41. doi: 10.1002/(sici)1097-0142(19991215)86:12<2632::aid-cncr7>3.0.co;2-a. PMID: 10594858.

16. Issa JP, Byrd JC. Decitabine in chronic leukemias. Semin Hematol. 2005 Jul;42(3 Suppl 2):S43–9. doi: 10.1053/j.seminhematol.2005.05.005. PMID: 16015505.

